# Spatiotemporal modelling of meteorological drivers and outbreak detection of Legionnaires’ disease in England

**DOI:** 10.64898/2026.08.03.26359560

**Authors:** Nyall Jamieson, Christiana Charalambous, David M. Schultz, Falguni Naik, David Howett, Gavin Dabrera, Ian Hall

## Abstract

Legionnaires’ disease is a severe respiratory illness caused by *Legionella* bacteria, with most cases occurring sporadically and environmental sources often unidentified. Effective outbreak detection requires understanding the spatiotemporal dynamics of sporadic cases and their environmental drivers. We developed a mechanistically informed spatiotemporal model integrating fine-scale spatial heterogeneity, multi-week meteorological influences, and extended temporal lags. The framework combines a negative binomial generalised additive model (GAM), a Besag–York–Mollié (BYM2) spatial component, and distributed lag nonlinear models (DLNMs) to capture nonlinear, delayed effects of temperature, dewpoint depression, precipitation, and cloud cover. These outputs generate a national daily index of weather-driven vulnerability, which is combined with hierarchical clustering to identify potential outbreaks. Across 2000–2019, our model improved outbreak detection in 15 of 20 years compared with the baseline UKHSA approach; in the remaining years performance was either equivalent (two years) or only slightly worse (three years, 0.98% reduction). Mean relative improvements were 6.0%, with a maximum of 14.4% in 2013. Improvements were consistent across months, and coarser 0.25° grid evaluations likely underestimate the model’s advantage at finer spatial scales. The analysis also clarified dual-stage Legionnaires’ disease dynamics, distinguishing environmental bacterial growth from the shorter infection window, and demonstrated the necessity of extended lags for accurate risk prediction. This framework provides a robust platform for targeted surveillance and predictive modelling, supporting evidence-based interventions and enhancing preparedness for sporadic Legionnaires’ disease under observed climatic conditions.

## 1 Introduction

Legionnaires’ disease is an illness resulting from infection by the gram-negative bacterium *Legionella* [1]. This bacterium proliferates in aqueous environments, including engineered systems such as cooling towers and plumbing networks, as well as natural aquatic habitats [1]. Through aerosolization, *Legionella* is dispersed into the air, where humans may inhale it. Following inhalation, the bacterium infects pulmonary tissue and triggers an inflammatory response [2]. Infected individuals typically develop pneumonia, which is required for a clinical diagnosis of Legionnaires’ disease, and may also experience symptoms such as fever, headache, and nausea. Severe complications can include respiratory failure, sepsis, and kidney failure, any of which can be fatal [1]. Because Legionnaires’ disease represents a substantial public health concern, authorities classify it as a notifiable disease in England [3]. Consequently, clinicians, laboratories, and other healthcare professionals are required to report all confirmed and suspected Legionnaires’ disease cases to their local proper officer or United Kingdom Health Security Agency (UKHSA) Health Protection Team (HPT), which in turn notifies the UKHSA National *Legionella* Surveillance Team [3].

Between 2000–2019, public health surveillance in England documented over 3,200 community-acquired Legionnaires’ disease cases [4]. These cases were classified as either sporadic or outbreak-associated based on temporal and spatial links [3]. Sporadic cases lacked identifiable epidemiological links, whereas outbreak cases involved individuals sharing microbiological or epidemiological connections, with symptom onset within 28 days of each other. Community-acquired clusters are defined as two or more cases occurring within six km and six months [3]. During 2000–2019, 78% of the reported cases of Legionnaires’ disease occurred sporadically, most without a known exposure source [5, 6]. Because sporadic cases rarely lead to source identification, public health efforts have placed particular importance on detecting clusters to identify high-risk sources (i.e., sources whose characteristics and environmental conditions, capacity to generate aerosols, and the number of people potentially exposed could facilitate *Legionella* transmission) [45]. These sources are sites where *Legionella* can proliferate to levels capable of causing widespread illness and mortality in a local population, highlighting the importance of cluster detection for preventing and identifying outbreaks.

Historical outbreaks provide additional insight into Legionnaires’ disease transmission in both urban and hospital settings. For instance, in April 1985, a Stafford hospital outbreak resulted in 68 confirmed cases of Legionnaires’ disease [7, 8]. In September–October 1988, two outbreaks in Bolton city center resulted in 58 confirmed cases of Legionnaires’ disease [9]. In January–February 1989, a cluster in London’s Piccadilly Circus involved 33 confirmed and nine suspected cases [10]. Barrow experienced a major outbreak in July–August 2002, which resulted in 179 confirmed cases of Legionnaires’ disease [11]. Additional outbreaks include Southeast London (14 cases, 2005) [12], Glasgow (33 cases, 1984) [13], Stoke-on-Trent (21 cases, 2012) [14, 15], and Edinburgh (50 cases, 2012) [16, 17]. Collectively, these events illustrated the heterogeneity of Legionnaires’ disease outbreaks in terms of magnitude, duration, and geographical setting. Therefore, adaptable detection strategies to identify outbreaks are essential. To address this heterogeneity issue, UKHSA implemented an outbreak detection algorithm that compared new-case reports against baseline sporadic distributions and flagged deviations using a hierarchical clustering algorithm to detect potential outbreaks [18]. This system required mathematical modelling to quantify baseline incidence rates and map the spatiotemporal distribution of sporadic cases at the postcode sector level [18]. Therefore, understanding the distribution of sporadic cases was also essential, as their incidence reflects underlying environmental and behavioral drivers that shape *Legionella* exposure risk.

Legionnaires’ disease cases are influenced by both environmental and human factors that affect bacterial growth, aerosolization, and inhalation risk [19, 20, 21]. In particular, four broad categories of environmental drivers have been emphasized in the literature: water temperature, relative humidity, ultraviolet (UV) radiation, and precipitation. Water temperatures between approximately 20–45°C *accelerate Legionella* proliferation within engineered and natural water systems. Heavy precipitation or flooding can lead to water accumulation in low-lying or poorly drained areas; following rainfall cessation, such environments may form stagnant pools that facilitate microbial amplification [22]. Once aerosolized, *Legionella* bacteria remain viable for longer periods under moderate relative humidity and low solar radiation, although the relationship between relative humidity and bacterial survival is non-monotonic [23, 24, 25, 26, 27]. Human behaviour further modulates exposure risk: increased use of cooling towers, spa pools, and showers during warm periods elevates inhalation risk, while plumbing stagnation during low-occupancy intervals increases bacterial concentrations [28].

A substantial body of epidemiological research has examined associations between meteorological conditions and Legionnaires’ disease incidence using statistical and mathematical models. Collectively, these studies span a wide range of temporal resolutions, meteorological predictors, lag structures, modelling frameworks, and spatial assumptions, reflecting both the ecological complexity of Legionnaires’ disease transmission and the lack of methodological consensus in the field, as summarized in a prior review [59]. The synthesis presented here draws on a diverse body of literature to characterise prevailing approaches and identify common limitations. The studies were identified through a search of papers that employed statistical models to examine the relationship between meteorological conditions and Legionnaires’ disease.

One major axis of variation across prior analyses concerns temporal aggregation. Researchers have modelled daily [6, 20, 29, 32, 33, 34, 38, 60, 61, 70], weekly [19, 21, 62], monthly [5, 20, 30, 31, 63], and yearly [64] incidence data (Table 1). Daily resolution aligns closely with incubation periods and short-term environmental variability but is sensitive to reporting noise and sparse counts. Weekly aggregation smooths stochastic variation and may better capture delayed effects, although it can dilute associations driven by extreme weather events. Monthly data provide greater statistical stability but risk obscuring short-term mechanistic relationships, while yearly aggregation is generally too coarse to capture biological drivers, reflecting only long-term climatic patterns.

**Table 1:** Summary of analyses on meteorological effect on Legionnaires’ disease incidence. Here, DLM represents distribution lag linear model, CLR represents conditional logistic regression, LR represents logistic regression, and PCA represents principal component analysis. Further, * indicates that the model attempted to estimate the infection date from the symptom-onset data and incubation-period model. *Positive* and *negative* denote the direction of association as reported by the original authors for at least one exposure contrast or lag period, and do not imply monotonic or causal effects.

| Paper | Reported direction of association | Temporal resolution | Lag | Method |
| --- | --- | --- | --- | --- |
| [31] | Precipitation: positive<br>Relative humidity: positive<br>Temperature: positive | Monthly | 14-day average | Poisson GLM |
| [32] | Precipitation: positive<br>Relative humidity: positive<br>Temperature: negative | Daily | 2–10 day | Poisson GLM |
| [38] | Relative humidity: direction varies by lag<br>Temperature: direction varies by lag<br>Wind speed: direction varies by lag | Daily | 2–10 day | DLNM |
| [70] | Precipitation: direction varies by lag<br>Relative humidity: direction varies by lag | Daily | NA* | DLM |
| [21] | Precipitation: positive<br>Pressure: mixed across lags<br>Temperature: mixed across lags | Weekly | 1–4 week | Poisson GLM |
| [62] | Cloudiness: mixed across lags<br>Precipitation: positive<br>Pressure: mixed across lags<br>Relative humidity: mixed across lags<br>Sunshine: mixed across lags<br>Temperature: positive | Weekly | 0 | Poisson GLM |
| [60] | Precipitation: positive<br>Relative humidity: mixed across lags<br>Temperature: negative | Daily | 0–15 day | CLR |
| [63] | Precipitation: mixed across lags<br>Pressure: positive<br>Relative humidity: positive<br>Temperature: positive<br>Wind speed: mixed across lags | Monthly | 0 | Poisson GLM |
| [34] | Relative humidity: positive<br>Temperature: positive<br>Wind speed: positive | Daily | 9-day moving average | Poisson GLM |
| [29] | Precipitation: positive<br>Pressure: mixed across lags<br>Relative humidity: positive<br>Temperature: positive<br>Wind speed: negative | Daily | 0–15 day | Poisson GLM<br>CLR |
| [6] | Dewpoint: mixed across lags<br>Precipitation: positive<br>Pressure: mixed across lags<br>Relative humidity: positive<br>Temperature: negative<br>Visibility: mixed across lags<br>Wind speed: negative | Daily | 0–15 day | Poisson GLM<br>CLR |
| [61] | Precipitation: positive<br>Relative humidity: positive<br>Temperature: positive<br>UV radiation: negative<br>Wind speed: negative | Daily | 2–10 day | CLR |
| [64] | Precipitation: positive<br>Sunshine: negative<br>Temperature: positive<br>UV radiation: negative | Yearly | 0 | Correlation |
| [5] | Precipitation: positive<br>Temperature: positive | Monthly | 0 | NG GLM |
| [19] | Cloudiness: negative<br>Precipitation: positive<br>Relative humidity: positive<br>Sunshine: negative<br>Temperature: negative<br>Wind Speed: positive | Weekly | 0–2 week | Poisson GLM<br>PCA |
| [33] | Precipitation: positive<br>Pressure: mixed across lags<br>Relative humidity: positive | Daily | 0–35 day | NB GLM<br>CLR |
| [20] | Precipitation: positive<br>Relative humidity: positive<br>Temperature: negative<br>Wind speed: negative | Daily<br>Monthly | 0–14 day<br>4–24 week | CLR |
| [30] | Precipitation: positive<br>Relative humidity: positive<br>Temperature: mixed across lags | Monthly | 0 | LR |

Across studies, meteorological predictors most frequently include temperature [5, 6, 19, 20, 21, 29, 30, 31, 32, 34, 38, 60, 61, 62, 63, 64] and precipitation [5, 6, 19, 20, 21, 29, 30, 31, 32, 33, 60, 61, 62, 63, 64, 70], followed by relative humidity [6, 19, 20, 29, 30, 31, 32, 33, 34, 38, 60, 61, 62, 63, 70], wind speed [6, 19, 20, 29, 34, 38, 61, 63], atmospheric pressure [6, 21, 29, 33, 62, 63], sunlight duration [19, 62, 64], ultraviolet radiation [61, 64], cloudiness [19, 62], visibility [6] and dewpoint temperature [6]. As summarised in Table 1, the reported direction of association for these variables varies substantially across studies and, in several cases, across lag windows within the same study. These variables capture distinct stages of the infection pathway, from bacterial growth and persistence in environmental reservoirs to aerosol transport and human exposure. For example, precipitation may influence aerosol washout [65], temperature governs both bacterial proliferation and seasonal human activity [66], relative humidity affects aerosol stability [25, 26, 27], and wind speed modulates dispersion and dilution of contaminated aerosols [67]. Ultraviolet radiation contributes to bacterial inactivation, while cloudiness and solar exposure reflect both physical and behavioural seasonal patterns [23, 24, 68].

Methodologically, most studies employ Poisson or negative binomial generalized linear models to analyse count outcomes [5, 6, 19, 21, 29, 31, 32, 33, 34, 62, 63], with modelling choices and lag specifications varying widely across analyses (Table 1). Seasonality is typically modelled using sine or cosine terms or calendar splines, implicitly assuming fixed cyclical patterns with constant peak timing and magnitude across years [29, 33, 34, 70]. Conditional logistic regression within case–crossover designs has also been widely used to control for time-invariant confounding [6, 20, 29, 33, 60, 61]. Only one study has applied a distributed lag nonlinear model (DLNM) to Legionnaires’ disease incidence [38], restricting analysis to lags of 2–10 days surrounding symptom onset.

Despite their widespread application, these approaches share several important limitations. Several models assume linear or quadratic relationships between meteorological variables and disease incidence [6, 31, 19, 32], failing to capture threshold effects or non-monotonic responses that are biologically plausible for *Legionella* ecology. Highly correlated predictors such as temperature and relative humidity are often included simultaneously, leading to unstable coefficient estimates. Temporal trends are frequently modelled using rigid sinusoidal terms that ignore interannual variability in peak incidence [29, 33, 34, 70]. Lag structures are commonly selected based on statistical significance for individual days [6, 32, 38, 61], even though meteorological conditions are likely to influence bacterial growth, aerosolization, and exposure cumulatively over multiple weeks. While DLNMs permit modelling of delayed effects, previous applications have focused on short lag windows corresponding only to the incubation period, neglecting earlier environmental conditions required for bacterial amplification [38].

Spatial exposure assessment presents an additional challenge. Most studies assign meteorological variables at the level of cities, regions, or weather stations, implicitly assuming spatially uniform exposure [19, 20, 21, 60, 61, 62, 63]. In contrast, Legionnaires’ disease is typically hyper-localized, associated with specific point sources such as cooling towers or water systems that respond heterogeneously to weather and local microclimates. Aggregation over large spatial units risks exposure misclassification and ecological fallacy, diluting true associations. Moreover, meteorological variables may influence disease incidence indirectly through their effects on environmental *Legionella* proliferation rather than acting as direct causal drivers, implying that observed relationships are unlikely to be strictly additive or monotonic [25, 26, 27].

Collectively, these methodological inconsistencies and simplifying assumptions have contributed to heterogeneous and sometimes contradictory findings across studies regarding the direction and magnitude of meteorological effects on Legionnaires’ disease incidence, as summarised in Table 1 [5, 20, 21, 29, 30]. They also limit the generalizability and predictive utility of existing models, particularly for applications such as outbreak detection. In addition, the current UKHSA outbreak detection algorithm assumes that meteorological measurements from a single reference location adequately represent conditions across England, neglecting spatial heterogeneity and local environmental variability known to influence *Legionella* ecology [18]. These limitations motivate the need for modelling frameworks that more realistically represent nonlinear, delayed, and spatially heterogeneous meteorological influences on sporadic Legionnaires’ disease risk.

To address these limitations and extend prior research, we implement a structured modelling framework that proceeds in three steps and culminates in outbreak detection. First, we develop a spatial model of sporadic Legionnaires’ disease incidence using a Poisson Besag–York–Mollié model (BYM2) [35], a scaled reparameterization of the classic BYM model [36]. Unlike the original BYM formulation, BYM2 resolves identifiability issues between the spatially structured and unstructured random effects by scaling both components to unit marginal variance and introducing a mixing parameter that governs their relative contribution. This improves interpretability of spatial heterogeneity and enables more stable estimation of persistent regional variation in case occurrence between 2000–2019. Second, we re-evaluate associations between meteorological variables and Legionnaires’ disease incidence using uni- and multivariate distributed lag nonlinear models (DLNMs) [37] within a case-crossover design. Unlike previous applications restricted to short lag windows [38], our models quantify meteorological effects over extended lags of 0–60 days, reflecting both the incubation period and the environmental growth phase that enables bacterial amplification. Third, we combine spatial probabilities with DLNM-derived weather offsets to construct a national-scale time-series model of sporadic Legionnaires’ disease risk, producing a daily index of weather-driven vulnerability. Finally, outputs from the spatial and national models are integrated into a hierarchical clustering algorithm adapted from the prior outbreak detection approach [18], with the aim of improving detection reliability and predictive capacity for environmentally mediated outbreaks. This approach may further enhance sensitivity and specificity relative to earlier methods, thereby supporting data-driven public health interventions for environmentally mediated pathogens.

## 2 Dataset and Methods

In this section, we describe the dataset used in the study and the methods used for modelling the spatiotemporal distribution of sporadic cases, as well as the outbreak-detection tool. Section 2.1 provides details of the dataset, including weather, community-acquired cases of Legionnaires’ disease, population, Index of Multiple Deprivation (IMD), and urbanisation data. The weather data is provided on a 0.25°× 0.25° latitude–longitude grid, whereas the remaining parts of the data are defined at the Lower Layer Super Output Area (LSOA) level [42]. The LSOA geography is a standard small-area geographic unit in England used for statistical reporting. Each LSOA comprises between 400–1200 households, with a typical resident population of 1000–3000 individuals. Following this, we dssiscuss the spatial scale at which the data is aggregated. Following this, section 2.3 outlines the previous outbreak-detection model [18] and introduces our updated version, along with the background spatiotemporal models to support each outbreak-detection model.

### 2.1 Dataset

We begin by providing an outline of the weather data and the reported Legionnaires’ disease case data. Next, we describe the spatial data, including population, sex ratio, proportion of the population aged *≥* 65 years, IMD, and the urban–rural classification. Finally, we describe the spatial scale for this analysis.

#### 2.1.1 Weather data

We use meteorological data from the ERA5 reanalysis, the fifth-generation global climate reanalysis produced by the European Centre for Medium-Range Weather Forecasts (ECMWF) [43]. ERA5 provides a physically consistent reconstruction of past atmospheric conditions by assimilating observational data into a numerical weather prediction model. The data spans 1940 to the present with hourly estimates on a 0.25°× 0.25° latitude–longitude grid.

From ERA5, we extract variables relevant to the environmental conditions that may influence *Legionella* ecology, transmission, and case reporting. Specifically, we use wind speed (m s ^−1^), 2m air temperature (K), 2m dewpoint depression (K), total cloud cover (proportion) and total precipitation (m). The dewpoint depression is the difference between the air temperature and the dewpoint temperature. Variables such as precipitation and cloud cover capture atmospheric conditions that may affect microbial persistence and dispersal, whereas air temperature and dewpoint depression may directly influence bacterial viability. All variables are provided hourly, and we aggregate these to daily means (or accumulations, as appropriate) for analysis.

In the present study, we use these meteorological variables in two distinct ways. First, for each region of England, we calculate the mean of all daily values between 2000–2019 and use these in the spatial model, which aggregates data over the entire period. This approach allows us to identify regions that consistently experience different weather patterns and to determine whether spatial variations in incidence may be related to long-term climatic differences across the country. Second, we incorporate these variables in a regression model to assess how short- to medium-term weather variations (0–60 days prior to a case) influence the risk of Legionnaires’ disease, while controlling for location. Specifically, the regression approach uses a DLNM, a framework that uses splines across both exposure values and time lags to account for nonlinear effects and delayed responses. Although the clinical incubation period for Legionnaires’ disease is typically 2–10 days, we extend the lag period to 60 days to capture not only the infection window but also potential longer-term environmental processes, such as *Legionella* proliferation and survival dynamics, that may precede human exposure. In this DLNM analysis, weather variables are expressed as deviations from the expected mean for the corresponding week of the year and location. This approach allows the model to quantify the effect of short- to medium-term departures from the seasonal trend on Legionnaires’ disease case occurrence, rather than the effect of the regular seasonal pattern itself. By isolating these deviations, we reduce confounding arising from seasonality, enabling a clearer assessment of independent meteorological effects. Furthermore, this formulation ensures that subsequent time-series models of seasonal trends do not redundantly account for variation already captured in the DLNM, maintaining a consistent and non-overlapping representation of seasonal effects.

#### 2.1.2 Legionnaires’ disease case data

We use microbiologically confirmed Legionnaires’ disease case data in England from 2000–2019, provided by UKHSA, where the national surveillance scheme for Legionnaires’ disease is managed. Each record corresponded to a single diagnosed patient who was reported to have clinical and/or radiological evidence of pneumonia. For each case, the data included the date of symptom onset, the LSOA of the individual’s home address, whether the case was outbreak-associated or sporadic, and the classification of exposure. Patients were also surveyed about their location history during the 10 days prior to symptom onset to cover the likely incubation period, including nights spent at home, traveling within the UK, traveling abroad, or in hospital (nosocomial), in accordance with the national surveillance scheme guidelines [44]. Outbreak cases are defined as those linked as part of a cluster of infected cases close in time and space (i.e., within 28 days and six km apart), whereas sporadic cases are not linked to any other confirmed case [3, 45].

Following completion and review of the national Legionnaires’ disease enhanced surveillance form, cases are categorized by source if one is identified but mostly by the most likely assumed source based on the primary type of exposure: community-acquired, travel within the UK, travel abroad, and nosocomial [3]. Individuals with one or more days of hospitalization or travel are excluded from the community-acquired case definition [3]. This classification, together with the outbreak/sporadic distinction, allows us to identify baseline sporadic, community-acquired cases for modelling.

#### 2.1.3 Population

We use mid-year population estimates for each year between 2000–2019 [46, 47], reported at the LSOA level. However, LSOA boundaries are not fixed: they are revised following each national census to reflect population change [48]. As a result, the definition of LSOAs differs across the study period. For example, population data for 2011–2019 use the 2021 LSOA estimates [49], whereas data for 2000–2010 use the 2011 LSOA estimates [50].

For each LSOA, the dataset includes total population, as well as stratifications by age and gender. In the 2000–2010 data, population counts are provided by single year of age (e.g., one year old, two years old, etc.), whereas in the 2011–2019 data, counts are reported in age intervals (0–15, 16–29, 30–44, 45–64, 65+). To maintain consistency across years, we aggregate the single-year data into age bins matching the later interval structure. Within this standardized age distribution, we treat the proportion of individuals aged 65 and older as an at-risk population [51], which is included as a covariate in the analysis.

#### 2.1.4 IMD

The IMD is a United Kingdom (UK) measure of relative deprivation calculated for each LSOA. This metric ranks areas by deprivation to identify disadvantaged communities and target interventions. This metric comprises seven domains with different relative contributions (shown in parentheses) [52]: the proportion of the population experiencing low income (22.5%), the proportion of working-age people who are involuntarily excluded from work (22.5%), low educational attainment and lack of skills (13.5%), the risk of premature death or impaired quality of life due to poor health (13.5%), the risk of personal and material victimisation (9.3%), physical and financial access to housing and local services (9.3%), and the quality of housing and the outdoor environment (9.3%). More deprived areas often have higher disease risk due to environmental or healthcare access factors [53].

Similar to the population data, the IMD data is not consistent between 2000–2019. For example, IMD data in England has three versions since 2000: 2010 [54], 2015 [55], and 2019 [56]. Government guidance recommends specific usage of these three versions of the data across years 2000–2019 [57]. Precisely, usage of the 2010 data is advised for the years 2000–2010. Usage of the 2015 data is advised for the years 2011–2015. Usage of the 2019 data is advised for the years 2016–2019. We follow the government guidance as we provide values for each cell and year to be used within the models. We use this cleaned version of the IMD as a covariate in the spatial distribution model.

#### 2.1.5 Urbanisation

We use binary data indicating whether an LSOA is urban or rural [58]. This data is defined as the 2021 classification, which follows the 2021 LSOA structure [49]. We incorporate this proportion as a covariate in the spatial distribution model, alongside the sex ratio, old age proportion, and IMD data.

#### 2.1.6 Areal unit approach and data merging

The spatial scale at which analyses are conducted is crucial. For example, aggregating to large administrative geographies (e.g., northwest England, northeast England, Yorkshire and the Humber, West Midlands, East Midlands, East of England, southwest England, southwest England, and London) can obscure within-region patterns. Relationships observed at this coarse scale may not hold at finer resolutions. Specifically, a region may appear low risk overall while containing localized high-risk pockets. Similarly, large grid cells may smooth away subtle shifts in incidence over time.

At the opposite extreme, using small units, such as LSOAs, can introduce new challenges due to the low case counts and sparse observations. Sparse case counts can create the appearance of spurious heterogeneity, where a few cases make one small area appear disproportionately high risk. Small numbers of observations also increase model uncertainty, which can make statistical inference unstable and reduce the reliability of model outputs.

Therefore, we adopt a compromise: a square-lattice structure of 0.25° × 0.25° latitude–longitude grids. This resolution aligns with the grid structure of the weather data from the DLNM (discussed in Section 2.1.1), ensuring compatibility across the components of our data.

This choice also minimizes the influence of the modifiable areal unit problem—a statistical phenomenon whereby analytical results depend on the size, shape, or boundaries of the regions used. Units that are too coarse can mask localized high-incidence areas, whereas units that are too fine can inflate apparent heterogeneity due to sparse data. By selecting 0.25° × 0.25° latitude–longitude cells, we reduce these risks while retaining sufficient spatial resolution to capture meaningful patterns in Legionnaires’ disease incidence.

The grid cells vary slightly in geographic area, particularly near the coast and due to the convergence of meridians toward the poles. However, this variation does not affect incidence calculations because rates are expressed per capita within each cell. Likewise, small differences in cell size do not bias spatial patterns of aggregated variables such as population, proportion aged *≥* 65, sex ratio, IMD, and urban–rural classification, as these are measured per capita or area-weighted within cells. Therefore, the chosen grid scale provides a consistent and interpretable framework for comparing the risk of Legionnaires’ disease across England.

For the reported Legionnaires’ disease spatiotemporal data, we aggregate cases using this defined square-lattice structure, with each cell containing the total number of reported cases from all LSOAs that fall within it. LSOAs that overlap multiple cells are assigned to the cell containing the largest proportion of their area, preserving the overall spatial distribution. We focus on sporadic, community-acquired cases to develop a spatiotemporal model representing baseline observed incidence, which is then used to predict outbreak events in the outbreak-detection analysis.

Population data are converted to the grid structure in a similar manner: each LSOA is assigned to a cell, and populations within the same cell are summed to yield a total cell population. This cell-level population serves as an offset in the spatiotemporal model, accounting for the larger expected number of cases in more populous cells. We also derive two additional covariates per cell: sex ratio (male:female) and the proportion of the population aged *≥* 65, as incidence is known to be higher in males and older individuals [51].

IMD and urban–rural classification, reported at the LSOA level, are aggregated to the grid structure in the same way. For IMD, the mean of all LSOAs within a cell provides a cell-average value, whereas for urban–rural classification, each cell is assigned the proportion of LSOAs classified as urban. This approach ensures that all key demographic and contextual variables are aligned with the square-lattice structure, facilitating consistent, interpretable analyses of Legionnaires’ disease risk across England.

### 2.2 Mathematical modelling

Here, we develop an outbreak-detection framework extending the approach of [18], which identified spatiotemporal clustering of Legionnaires’ disease under an inhomogeneous Poisson assumption. In contrast to the Poisson formulation, our model represents sporadic, community-acquired cases as a stochastic count process following a negative binomial distribution, better capturing the observed variability in case counts. The framework further incorporates overdispersion, spatial heterogeneity, and meteorological effects, providing a more flexible and realistic representation of sporadic Legionnaires’ disease incidence. The original model of [18] is retained as a comparator to evaluate the performance of these extensions.

All analyses are conducted on the spatial grid structure of England defined in Section 2.1.1, which partitions the country into 347 regions indexed by *i∈* 1, … , 347. This grid provides the spatial framework for both the baseline incidence model and the outbreak-detection algorithm described below. Formally, we define

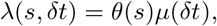

where *λ*(*s, δt*) represents the expected number of sporadic Legionnaires’ disease cases within a region *s* over the time period *δt* = *{t*_1_, … , *t*_*q*_*}* (corresponding to days *t*_1_ through *t*_*q*_).

We distinguish between two spatial indices, *i* and *s*, which serve different roles in the analysis. The index *i* denotes the fixed spatial units at which the underlying case data and covariates are defined, corresponding to the gridded weather-cell structure (*i* = 1, … , 347). All epidemiological observations and environmental covariates are initially available at this spatial resolution. In contrast, *s* does not represent a fixed spatial partition, but rather an analysis-level spatial construct. Specifically, *s* indexes arbitrary subregions of England, which in this study are defined as circular regions used in the case-pair outbreak analysis (see section 2.2.4). For each *s*, case counts are obtained by aggregating observations from the *i*-level grid cells that fall within the corresponding region. The use of both indices is therefore intentional: *i* represents the native spatial resolution of the data, while *s* represents the spatial scale at which outbreak characteristics are evaluated.

#### 2.2.1 Spatial component of the sporadic incidence model

We first focus on the spatial component, *θ*(*s*), which describes the proportion of cases expected within a region *s*. Unlike the previous model, which assumes uniform per-capita incidence, we allow *θ*(*s*) to vary according to demographic, socioeconomic, and environmental factors, including age distribution, sex ratio, deprivation, urbanisation, and the weather variables listed in Section 2.1.1. This formulation acknowledges that risk is not uniform: older populations, certain genders, and residents of urban or deprived areas may experience different exposure and infection risks.

To formalize this, we adopt a two-stage approach. In the first stage, we construct a BYM2 model of incidence across England, incorporating the aforementioned covariates and spatial autocorrelation. In the second stage, we use the central estimates of this model to generate a proportional map of expected cases for each region, ensuring that the total proportion sums to one.

Mathematically, let *Y*_*i*_ denote the total observed Legionnaires’ disease case count in spatial region *i* over the entire study period 2000–2019. With this, *i ∈ {* 1, … , 347 *}*and let *N*_*i*_ be the population at risk. We model

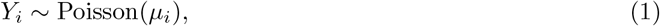

where *µ*_*i*_ is the expected case count for region *i*, conditional on covariates and spatial effects. For the 20-year aggregated data, the variance closely matches the mean, indicating no substantial overdispersion; hence, a Poisson specification suffices for the temporally aggregated dataset. We link the expected counts to covariates and population through

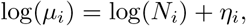

where the linear predictor *η*_*i*_ is given by

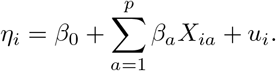

Here, *β*_0_ denotes the model intercept, *X*_*ia*_ denotes the covariates for region *i* (e.g., demographic, deprivation, urbanisation, and meteorological variables), *β*_*a*_ are the corresponding regression coefficients, and *u*_*i*_ is a spatial random effect that captures residual variation not explained by the covariates. We model *u*_*i*_ using the BYM2 specification, which separates spatial variation into two components: a structured component that reflects correlation between neighboring regions, and an unstructured component that accounts for independent, region-specific variation. In other words, we set:

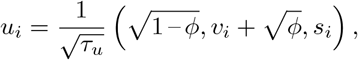

where *v*_*i*_ *∼ N* (0, 1) represents unstructured noise, *s*_*i*_ is the standardized structured spatial effect, *ϕ ∈* [0, 1] controls the proportion of variance attributable to the structured component, and *τ*_*u*_ is the overall precision. The structured effect *s*_*i*_ follows a conditional autoregressive (CAR) model:

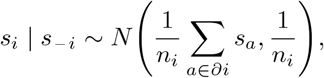

where *∂i* denotes the neighbors of region *i* and *n*_*i*_ = |*∂i*|. This formulation allows information to borrow strength across neighboring regions: if a region has few observed cases, its structured effect is informed by its neighbors, reducing noise and improving estimation. Standardization ensures that the structured and unstructured components are comparable, with total variance Var(*u*_*i*_) = 1*/τ*_*u*_. In practice, the BYM2 model provides a flexible way to capture both localized random fluctuations and broader spatial patterns, helping to identify areas with genuinely elevated risk while accounting for underlying spatial structure.

#### 2.2.2 Temporal component of the sporadic incidence model

Having established the spatial component, we next define the temporal component, *µ*(*δt*), representing the expected national incidence over *δt*. We express this as a sum of daily expected cases:

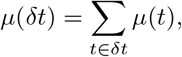

where *µ*(*t*) is the expected national incidence on day *t*. Daily incidence is modelled using a negative binomial (NB) GAM with mean *κ*(*t*) and dispersion parameter *ϕ*_1_. Unlike the temporally aggregated model *θ*(*s*), overdispersion is present in the spatially aggregated time series of national incidence. Additionally, we define the dispersion parameter *ϕ*_1_ to be independent of time for model parsimony; using a time-dependent parameter does not improve model fit. The GAM model is provided as follows:

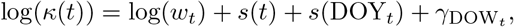

where *t* indexes calendar dates. The function DOY_*t*_ returns the day of the year corresponding to date *t*, which is an integer between 1–365, whereas DOW_*t*_ returns the day of the week corresponding to date *t*, which was an integer between 1–7. Therefore, *s*(DOY_*t*_) represents a smooth function on day of the year, and *γ*_DOW*t*_ represents a random effect on day of the week. Further, we included a random intercept for day of the week, modelled as a normally distributed effect with mean zero and estimated variance, to account for systematic differences in healthcare-seeking behavior or reporting to UKHSA across weekdays.

The term *s*(DOY_*t*_) is included as a proxy to capture the combined effects of seasonal environmental conditions (e.g., higher temperatures in summer) and seasonal human behaviour (e.g., prolonged holidays in summer leading to reduced water use in buildings, which may promote bacterial growth). Further, *w*_*t*_ represents a weather-related offset, capturing the influence of meteorological conditions on the likelihood of case occurrence in the days preceding the case. For clarity, we distinguish *w*_*t*_ from the weather proxy captured by *s*(DOY_*t*_): *s*(DOY_*t*_) reflects the average seasonal trends over the year, whereas *w*_*t*_, estimated from the DLNM, represents deviations from these average trends at a specific time (e.g., if September is warmer than usual, *w*_*t*_ quantifies the effect of this anomaly on incidence). This separation allows us to distinguish the overall seasonal pattern from short-term weather fluctuations. We consider both 0–30 and 0–60 days preceding a case to evaluate the importance of accounting for meteorological effects up to 60 days prior to infection. This offset is defined in equation (2).

Furthermore, we fitted a thin-plate regression spline on the date *t* (in days since start of series). A thin-plate regression spline captures nonlinear patterns, automatically selects knot placement, and includes a smoothing penalty to prevent overfitting by controlling the spline’s flexibility. This spline can reveal long-term trends and seasonal fluctuations without imposing strict assumptions. Additionally, this spline performed well at the boundaries compared with polynomial splines. Next, we placed a cyclic, cubic regression spline on day of the year to capture seasonal changes throughout each year and remove any edge effects at 31 December – 1 January.

For the splines used in this GAM, we selected 690 and 35 knots, respectively. Knot placement was guided by a general smoothing heuristic that links knot density to the characteristic timescale of disease development, corresponding to approximately one knot every two incubation-period cycles [71]. For Legionnaires’ disease, this timescale corresponds to 2 × 5.3 days, based on empirical incubation-period data [39]. Although this heuristic was originally proposed in the context of person-to-person viral infections, we apply it here as a pragmatic guideline for capturing short-term temporal structure rather than as a strict biological assumption.

To assess sensitivity to this choice, we also tested an alternative knot placement based on a 10-day incubation period, reflecting assumptions used in earlier UK Legionnaires’ disease studies [2, 72], corresponding to one knot every 2 × 10 days. Results were similar under both specifications, likely because the long time series provides sufficient flexibility such that inference is not sensitive to the precise knot interval at this level. We therefore adopted the 2 × 5.3-day knot spacing in the main analysis, as it is supported by empirical incubation-period data, whereas the alternative relies on earlier and less precise assumptions.

#### 2.2.3 Weather-related component of the sporadic incidence model

We now move onto the weather-related component of this model, which leads to a derivation of *w*_*t*_. To model short-term weather effects on Legionnaires’ disease incidence, we use a matched case-control conditional logistic regression design. This approach allows us to estimate how deviations (from the mean) in weather conditions influence the risk of disease while controlling for confounding factors such as region and time. Essentially, we quantify how short-term weather deviations influence Legionnaires’ disease risk, while controlling for confounding due to time (seasonality) and space (different regions may have different baseline risk). Matching each case with controls from the same region and week ensures that differences we see in weather effects are not due to underlying population or seasonal differences. A conditional logistic regression approach allows us to focus only on within-stratum comparisons, giving a clean estimate of the effect of weather deviations.

To begin, we again let *i* index regions and *t* index days in the time series. Each sporadic, community-acquired case of Legionnaires’ disease is assigned to a region *i* and a day *t*. For each case *k*, we define a case-control stratum *S*_*k*_ containing the case and up to *C*_*k*_ *≤*50 control observations from the same region *i* and the same week to form the dataset used for this case-crossover analysis. We organize the data so that each case is compared to similar control days in the same region and week. With this approach, we define the binary outcome variable

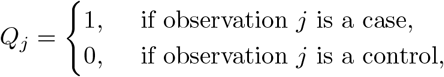

for each observation *j* in the dataset. This binary variable allows the regression model to estimate how weather changes the odds of being a case within the matched stratum. Next, let 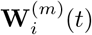 denote the exposure lagged for the weather variable *m* at lag *l*, where *t* = 0 corresponds to the day of the case. In our analysis, we include the mean values of air temperature, negative dewpoint depression, cloud cover, and precipitation. Including further weather variables (e.g., UV radiation, wind speed) leads to correlation issues, so a compromise is required. Air temperature, dewpoint depression and precipitation are crucial to any meteorological model, due to the known biological processes and the meteorologically related effects associated with these three variables. We included negative dewpoint depression (dewpoint temperature minus air temperature) to remove correlation between dewpoint temperature and air temperature. This formulation also ensures the variable (negative dewpoint depression) increases with relative humidity, which improves interpretability. Additionally, we include cloud cover as a covariate, serving as a practical proxy capturing potential tradeoffs with UV radiation in the model. With the chosen variables, lagged exposures up to *L* = 60 days are organized in a matrix

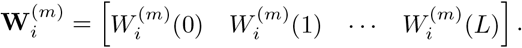

We choose such a long period of time preceding a case as the meteorological effects on Legionnaires’ disease are not instantaneous. By looking at lags up to 60 days, we capture both immediate effects (0–15 days before a case) and longer-term effects (e.g., conditions that allowed bacteria to grow and persist). Structuring this as a matrix lets us model all these lagged effects simultaneously. Following this, to model distributed-lag effects 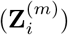 , we apply a natural cubic spline basis **B**(*t*) to the lag dimension. Weather effects over time are rarely linear. Therefore, splines let us smoothly model nonlinear patterns across the 60-day lag, so we can capture peaks and troughs of risk associated with short-term weather fluctuations. Internal and boundary knots specify where the spline can bend to fit the data. The basis has internal knots at 3, 7, 14, 35, 56 days and boundary knots at 0 and 60 days. This placement provides flexibility to capture variation within the infection window while imposing smoother behaviour across the longer bacterial-growth window. The distributed-lag effect is defined as:

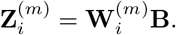

Concatenating the spline-transformed lag matrices for all exposures allows us to fit all weather effects together and account for their correlations. Concatenating all *p* exposure variables gives the full design matrix:

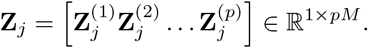

For each stratum *S*_*k*_, the conditional logistic regression model is

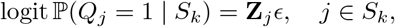

where 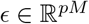 is the vector of regression coefficients for all lagged exposures. The model estimates how each weather variable at each lag affects the odds of being a case within the stratum. Conditioning on the stratum ensures we are comparing within the same region and same week, which removes confounding. The stratum-conditioned likelihood is:

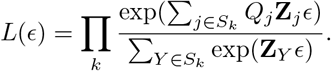

This likelihood ensures the probability of being a case is only evaluated relative to the matched controls, which is exactly what allows us to isolate the weather effect. Lag-specific log hazard ratios for exposure variable *j* at lag *l* are obtained as

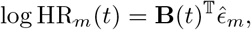

with standard errors

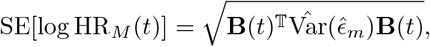

and corresponding confidence intervals

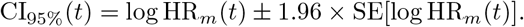

We translate the regression coefficients into lag-specific risk estimates, describing how much a weather variable at a particular lag influences risk. With this weather-deviation DLNM model, we develop a weather-related offset for the GAM model. To do this, we first consider all region–day pairs (*i, t*) in England and the time series 2000–2019, and match each pair to the nearest stratum. With *i* indexing regions and *t* indexing days in the study period, we construct the same lagged exposure representation and spline basis used for model fitting. After fitting the conditional logistic regression model, we generate predicted weather-related effects for every region–day pair (*i, t*) in the study period. For each weather variable, we construct lag matrices up to 60 days, using the same lag structure and natural cubic spline basis as in the fitted model. For prediction, each region–day pair is assigned to a case–control stratum. If a day naturally falls within a stratum (i.e., it was part of a matched case–control set), we use that stratum directly. Otherwise, we assign the region–day to the nearest stratum by minimizing a distance metric that combines temporal and spatial proximity:

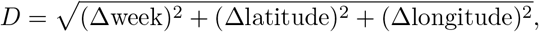

where Δweek is the cyclic difference in week numbers, and Δlatitude and Δlongitude are differences in stratum centroid coordinates. The region–day is then assigned to the stratum corresponding to the nearest case according to metric *D*. Predicted log odds for each region–day are obtained by multiplying the DLNM-transformed exposure vectors with the estimated regression coefficients from the fitted model.

To obtain a relative risk measure, we exponentiate these log odds and normalize them by the mean across all region–day pairs:

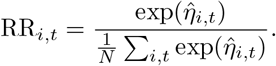

where *η*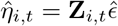 is the estimated log odds for region *i*, day *t*. Additionally, we define *N* as the number of region–day pairs in the dataset (i.e., *N* = (*i, t*) : |*i* = *i, .. .*, 347 and *t* = 1, *... J*| , where *J* is the number of days from 1 January 2000 to 31 December 2019). We convert the log odds from the regression into relative risk, scaled so that a value of one equates to expected risk at this location in this time of the year, which provides an intuitive measure. Spatial weights for each region are computed from the expected number of cases per region *µ*_*i*_ from the spatial model *θ*(*s*). Define the proportion of expected cases:

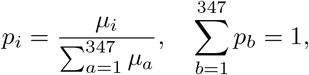

representing the relative contribution of region *i* to national cases. The national-scale, spatially weighted weather effect on day *t* is then:

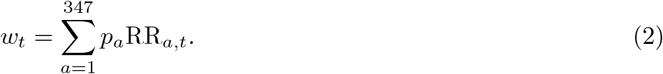

#### 2.2.4 Outbreak-detection algorithm

Finally, we introduce the outbreak-detection algorithm. The core idea is to quantify, for each ordered pair of cases, how likely it is that their observed spatiotemporal proximity could arise under the model of sporadic, community-acquired incidence. For each ordered pair of cases, we define a spatiotemporal window determined by the pair. The temporal component of this window, *δt*, is defined as the interval between the dates of symptom onset of the first and second cases. The spatial component of this window, *s*, is defined by constructing a circle centred at the location of the first case, with radius equal to the Euclidean distance between the two cases. By construction, the second case lies on the circumference of the resulting region *s*.

Given this spatiotemporal window, we estimate the number of sporadic cases expected under the baseline model. Let *X* denote the random variable representing the number of sporadic, community-acquired Legionnaires’ disease cases occurring within region *s* over the interval *δt*. Our goal is to estimate the probability *P* (*X >* 1), which quantifies the probability that at least one more sporadic case is expected within this spatiotemporal window. Because the joint distribution of *X* is non-standard, we estimate *P* (*X >* 1) using Monte Carlo simulation.

For each case pair, we first sample the total number of sporadic cases expected nationally over *δt* from:

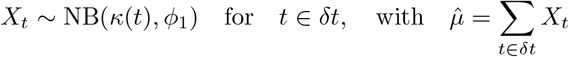

denoting the resulting draw. Conditional on this draw, the number of cases expected within spatial region *s* is sampled from a binomial distribution:

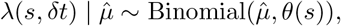

yielding a realization 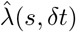. This two-stage sampling procedure is repeated 10,000 times for each ordered case pair, allowing us to estimate *P* (*X >* 1) empirically. Higher values of *P* (*X >* 1) indicate that observing a second case within the corresponding spatiotemporal window would be expected under sporadic incidence. In contrast, lower values indicate that the observed spatiotemporal proximity of the case pair is unlikely to have occurred by chance, providing evidence of potential outbreak-related clustering.

These probabilities are then used as a distance metric for hierarchical clustering with average linkage. Average linkage groups cases based on their proximity to the mean rather than extreme distances, producing compact clusters that are robust to outliers. Applying this procedure across all ordered case pairs enables the identification of meaningful spatiotemporal clusters while accounting for overdispersion, spatial heterogeneity, and weather-related effects.

## 3 Results

In this section, we present the results of fitting the various components of our outbreak-detection framework to the data set.

### 3.1 Spatial component of background sporadic model

Understanding the spatial distribution of Legionnaires’ disease is crucial for identifying regions of elevated risk and for generating hypotheses about the drivers of incidence. Spatial modelling enables us to account for both measured covariates and residual spatial dependence, thereby quantifying structured and unstructured variation in disease risk. This framework identifies areas with incidence elevated relative to the national average, which may reflect local infrastructure, population density, or environmental conditions conducive to *Legionella* proliferation. Together, these spatial patterns provide a basis for formulating hypotheses about how local environmental, demographic, and infrastructural factors shape observed incidence.

Parameter estimates from the Poisson BYM2 spatial model [equation (1)] fitted to the data are presented in Table 2. These model outputs describe associations between socioeconomic, demographic, and environmental variables and spatial variation in Legionnaires’ disease incidence.

**Table 2:** Summary of Poisson BYM2 model [equation (1)] outputs. Here, SD represents the standard deviation.

| Variable | Mean | SD | 2.5% percentile | Median | 97.5% percentile |
| --- | --- | --- | --- | --- | --- |
| (Intercept) | −11.747 | 1.890 | −15.461 | −11.744 | −8.047 |
| IMD | −0.060 | 0.029 | −0.118 | −0.060 | −0.002 |
| Sex ratio | 1.930 | 1.844 | −1.680 | 1.928 | 5.553 |
| Proportion of $\geq 65$ individuals | −0.261 | 1.496 | −3.194 | −0.261 | 2.675 |
| Wind speed | 0.110 | 0.119 | −0.123 | 0.110 | 0.345 |
| Negative dewpoint depression | −0.396 | 0.127 | −0.646 | −0.396 | −0.145 |
| Air temperature | 0.129 | 0.225 | −0.312 | 0.129 | 0.571 |
| Precipitation | −0.052 | 0.121 | −0.289 | −0.052 | 0.185 |
| Cloud cover | 0.257 | 0.216 | −0.167 | 0.257 | 0.682 |

We first considered socioeconomic and demographic influences. Higher IMD was associated with a small reduction in Legionnaires’ disease incidence (posterior mean = – 0.06 per unit increase), with the 95% credible interval narrowly excluding zero (Table 2). This association was weak indicating a modest association. The observed pattern may reflect spatial heterogeneity in environmental exposure that is not directly aligned with these indexes of multiple deprivation, although this interpretation is speculative.

Building on the discussion of IMD, sex-related effects on Legionnaires’ disease incidence were less consistent. Regions with a higher proportion of males tended to exhibit enhanced incidence (posterior mean = 1.930 per unit increase), although the wide 95% credible interval (– 1.68 to 5.55) indicated substantial uncertainty. This variability may have partly resulted from the spatial resolution of the gridded population data, which attenuated sex-related effects due to the approximately uniform gender distribution within each cell. At finer spatial resolutions, or when incorporating daily activity patterns that segregate populations by sex, stronger sex-related associations may become apparent.

Similarly, the proportion of older individuals exhibited no clear association with Legionnaires’ disease incidence (95% credible interval of – 3.19 to 2.68). This indicates limited evidence for a population-level age effect within the spatial model. Although older age is a known individual-level risk factor for severe disease, age measured as an area-level proportion may be an imprecise proxy for exposure risk and underlying susceptibility, which could contribute to the absence of a detectable association. One possible explanation is that, at the population level, older adults may have reduced contact with common environmental sources of *Legionella*, such as workplaces or recreational facilities, which could partially offset increased biological susceptibility; however, this interpretation remains speculative.

Next, we considered meteorological factors. Wind speed was weakly positively associated with Legionnaires’ disease incidence (posterior mean = 0.11), although the 95% credible interval (– 0.12 to 0.35) included zero. This interval indicates limited statistical support for an association between wind speed and incidence at the spatial scale considered. The positive posterior mean is nevertheless suggestive of a potential mechanism whereby moderate wind speeds may enhance the dispersion of aerosolized *Legionella*, increasing the spatial reach of exposure. At the same time, too low wind speeds may limit dispersion and reduce exposure probability, whereas high wind speeds could dilute bacterial concentrations, lowering the likelihood of sufficient inhalation at any given location. Such a pattern is consistent with a non-linear effect of wind on exposure, although our data do not directly quantify this relationship, and any conclusions should be interpreted cautiously.

Temperature-related variables showed clearer associations with Legionnaires’ disease incidence. Specifically, higher air temperature was associated with increased incidence (posterior mean = 0.13; 95% credible interval: – 0.31 to 0.57), whereas greater negative dewpoint depression was associated with reduced incidence (posterior mean = – 0.40; 95% credible interval: – 0.65 to – 0.15). These associations are consistent with the hypothesis that elevated temperatures promote *Legionella* proliferation in water systems, while conditions with lower dewpoint depression may reduce bacterial survival in aerosols. Moreover, human behavior may interact with these environmental conditions: warmer temperatures can increase the use of potential exposure sources such as spas, cooling towers, and showers. While these explanations are plausible and 95% credible intervals are wide, the data do not allow direct inference of mechanistic processes, and these interpretations should also be considered as tentative hypotheses suggested by the observed associations.

Greater cloud cover was associated with a modest increase in incidence (posterior mean = 0.26; 95% credible interval: – 0.17 to 0.68). Similarly, precipitation showed a small negative association (posterior mean = – 0.05; 95% credible interval: – 0.29 to 0.19). While these effects are weak and the 95% credible intervals include values close to zero, they may suggest environmental influences on *Legionella* survival and dispersal. For instance, cloud cover could indirectly affect bacterial persistence through reduced UV radiation, and light rainfall might facilitate aerosolization from environmental reservoirs, whereas heavy rainfall could dilute bacterial concentrations.

It is important to note that the spatial model aggregates case counts over the entire 2000–2019 period and uses long-term averages of population and meteorological variables. Consequently, this approach captures broad spatial associations between environmental factors and incidence, rather than day-to-day or short-term weather-driven fluctuations in case counts. Unlike time-series or distributed-lag analyses, the model does not allow for fine-grained assessment of the temporal relationship between weather and disease risk. Instead, the spatial effects primarily reflect regions where environmental conditions over the long term are broadly conducive to Legionnaires’ disease occurrence, potentially in combination with population density, infrastructure, and other unmeasured spatially structured factors. This framing helps interpret modest or weak associations between weather variables and incidence in the spatial model: they indicate patterns consistent with long-term suitability, rather than mechanistic causal effects of short-term weather variation.

Finally, to complement the demographic and meteorological analyses, we examined the spatial structure of Legionnaires’ disease risk using the BYM2 model parameters. The estimated precision of the spatial random effect was 5.70, corresponding to a variance of 0.18, while *ϕ* = 0.88. This estimate of *ϕ* indicates that approximately 88% of the spatial variation in Legionnaires’ disease incidence is attributable to structured effects, meaning that neighboring regions tend to exhibit similar incidence levels. Such spatial dependence may arise from shared environmental exposures, urban infrastructure, or local population behaviors not fully captured by the measured covariates. We examined spatial hotspots to better understand the distribution of sporadic, community-acquired Legionnaires’ disease (Figure 1).

**Figure 1.**
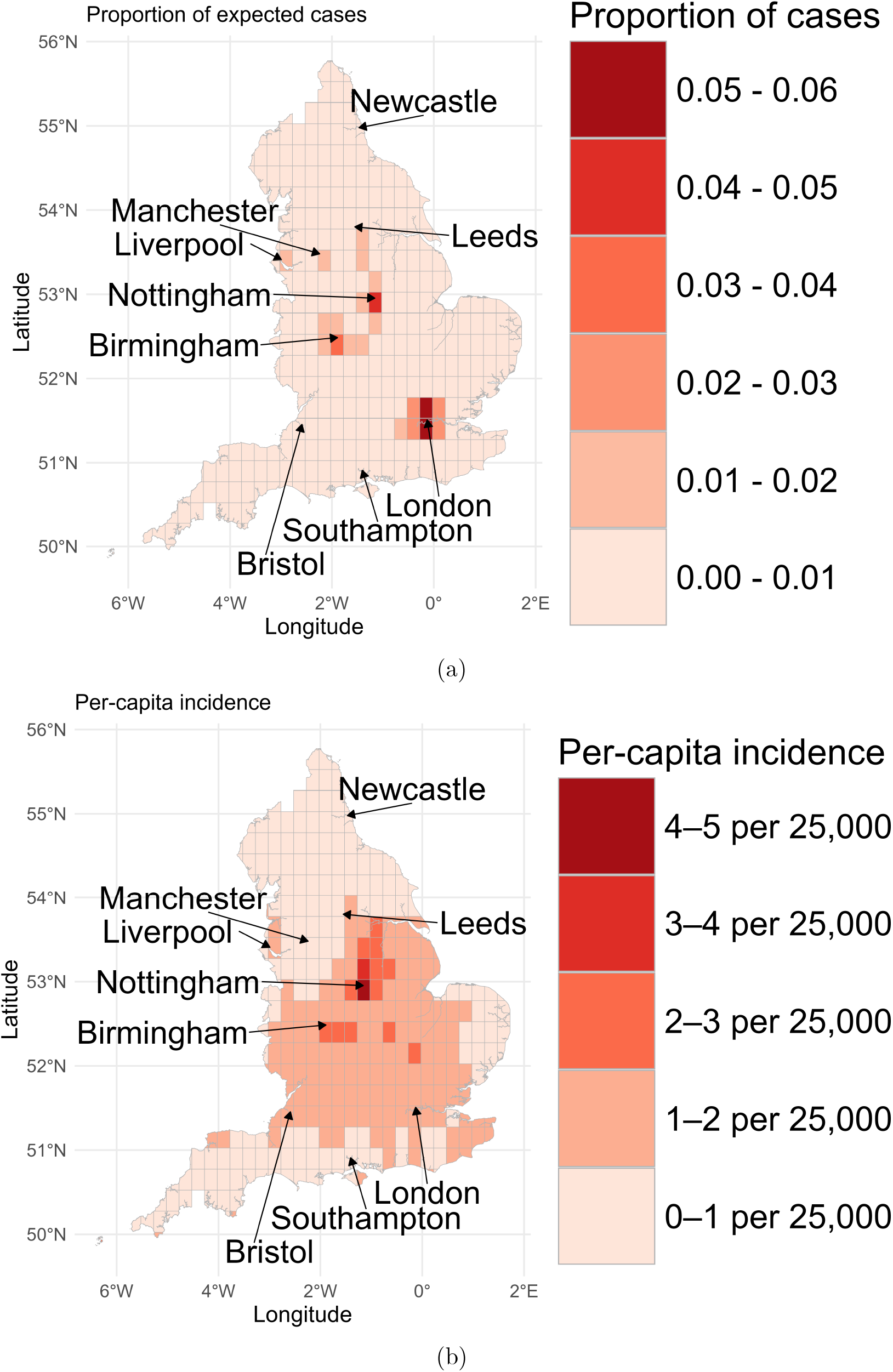
Plots of the spatial distribution of sporadic, community-acquired Legionnaires’ disease cases. Subfigure (a) illustrates the proportion of sporadic, community-acquired cases that occurred within each cell of the lattice-grid structure. Subfigure (b) illustrates the per-capita incidence estimated by the model.

Analysing cases as proportions of total incidence highlighted clustering in high-population regions, including London, Birmingham, Manchester, Liverpool, and Nottingham, where covariates and local population size reinforced elevated risk (Figure 1(a)). However, raw proportions obscured per-capita patterns, which became evident when examining the predicted incidence adjusted for population. Percapita incidence was particularly pronounced in Nottingham, the East and West Midlands, and areas extending toward London, whereas northern, southwestern, and eastern regions consistently exhibited lower incidence (Figure 1(b)). This correspondence between modelled spatial structure and observed incidence underscored the importance of considering regional clustering and local environmental or infrastructural features when interpreting Legionnaires’ disease dynamics.

### 3.2 Weather component of the background sporadic model

Understanding how deviations from typical seasonal weather influence Legionnaires’ disease incidence was critical for capturing the environmental drivers of transmission. Although seasonal averages of weather describe general trends, short-term anomalies (e.g., unusually warm, humid, or wet periods) can create conditions that either favor or inhibit bacterial growth and human exposure. The DLNM model allowed us to quantify these lagged, nonlinear effects, linking weather conditions preceding a case to the relative risk of disease. This approach provided insight into the temporal windows during which meteorological factors are influential, improving both mechanistic understanding and predictive modelling of outbreaks (Figure 2).

**Figure 2.**
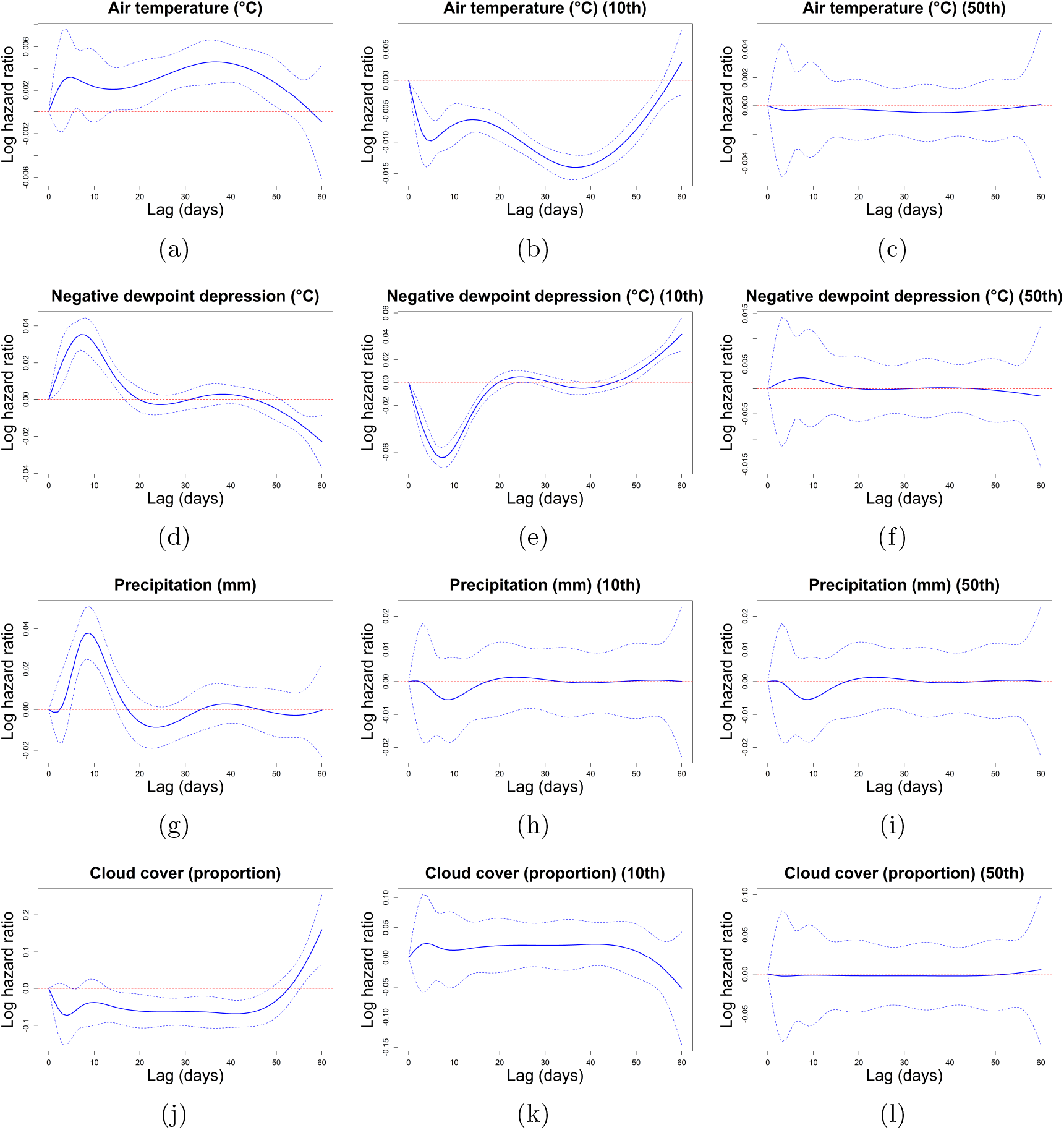
A plot of the estimated effects of deviations from the seasonally controlled mean of meteorological variables on Legionnaires’ disease incidence. Subfigures (a–c) represent air temperature effects. Subfigures (d–f) represent the negative dewpoint depression effect. Subfigures (g–i) represent the precipitation effect. Subfigures (j–l) represent the cloud cover effect. Subfigures (a, d, g, j) represent the effect of a one-unit increase from zero. Subfigures (b, e, h, k) represent the effect of one-unit increase from the 10^th^ quantile. Subfigures (c, f, i, l) represent the effect of a one-unit increase from the median.

Temperature deviations from the seasonal expectation exhibited a bimodal association with Legionnaires’ disease incidence (Figure 2(a)). In the 0–15-day lag window, higher-than-expected temperatures were associated with elevated risk, whereas a second smaller increase in risk occurred at 25–50-day lags. At the 10th percentile of seasonal temperature (*≈* 3°C below expected), a 1°C increase was associated with reduced risk (Figure 2(b)), while at the median, wide confidence intervals indicated substantial uncertainty (Figure 2(c)). One potential explanation for the bimodal pattern is that short-term warm periods may coincide with increased human contact with water systems or environments colonized by *Legionella*, while longer lags may reflect time for bacterial replication and accumulation in environmental sources, although this mechanism is speculative and not directly tested in the current analysis.

Building on this understanding of temperature effects, we next evaluated the role of negative dewpoint depression deviations. Negative dewpoint depression deviations from the seasonal expectation exhibited a nonlinear association with Legionnaires’ disease incidence within the infection window (Figure 2(d)). At the 10th percentile (*≈* – 1.8°C below expected), a 1°C increase was associated with a reduction in risk (Figure 2(e)), whereas at the mean level (0°C deviation), increases corresponded to elevated risk (Figure 2(d)), highlighting that the effect of dewpoint depression deviations within the infection window are inherently nonlinear. Outside the infection window, the model indicated minimal or no association between negative dewpoint depression deviations and incidence, suggesting that the influence of dewpoint depression is restricted to the infection window. One possible explanation is that deviations in the negative dewpoint depression during the infection window have a nonlinear effect on bacterial survival in water systems or aerosols, which is consistent with known biological experiments [25, 26, 27].

In addition to temperature and dewpoint depression, precipitation patterns further influenced *Legionella* dynamics. Precipitation deviations from seasonal expectations also exhibited a nonlinear relationship with Legionnaires’ disease incidence within the infection window (Figure 2(g)). At the lower end of the distribution (*≈* 10th percentile, – 0.14 mm below expected), increases in precipitation were associated with slight reductions in risk (Figure 2(h)), whereas higher-than-average precipitation was linked to modestly increased incidence (Figure 2(g, i)). These contrasting effects further illustrate the nonlinear influence of rainfall on disease risk. Outside the infection window, precipitation deviations had minimal apparent effect on incidence, indicating that the timing of rainfall relative to infection is critical. A possible, speculative interpretation is that light precipitation may transiently remove bacteria from environmental sources or dilute aerosols, while heavier rainfall could promote human exposure to indoor water systems (e.g., showers, taps) where bacteria persist.

Cloud cover deviations from seasonal expectations showed a subtle and nonlinear association with Legionnaires’ disease incidence (Figure 2(j)). Lower-than-expected cloud cover (*≈* 10th percentile, 32% below expected) was associated with slightly increased risk (Figure 2(k)), whereas near-median deviations had minimal effect, and the confidence intervals were wide (Figure 2(l)). This pattern highlights both the uncertainty of the estimate and the potential nonlinear influence of cloud cover on disease risk. A speculative explanation is that reduced cloud cover may increase UV exposure, potentially impacting bacterial survival in outdoor sources or modulating human behaviors that influence exposure, though these mechanisms were not directly measured. Overall, these results suggest that cloud cover deviations have at most a modest influence on Legionnaires’ disease incidence within the infection window.

Following this analysis, we quantified how meteorological conditions from 2000–2019 influenced Legionnaires’ disease incidence, capturing both within-year seasonal effects and between-year variation in factors affecting bacterial growth, aerosolisation, survival, and human exposure (Figure 3).

**Figure 3.**
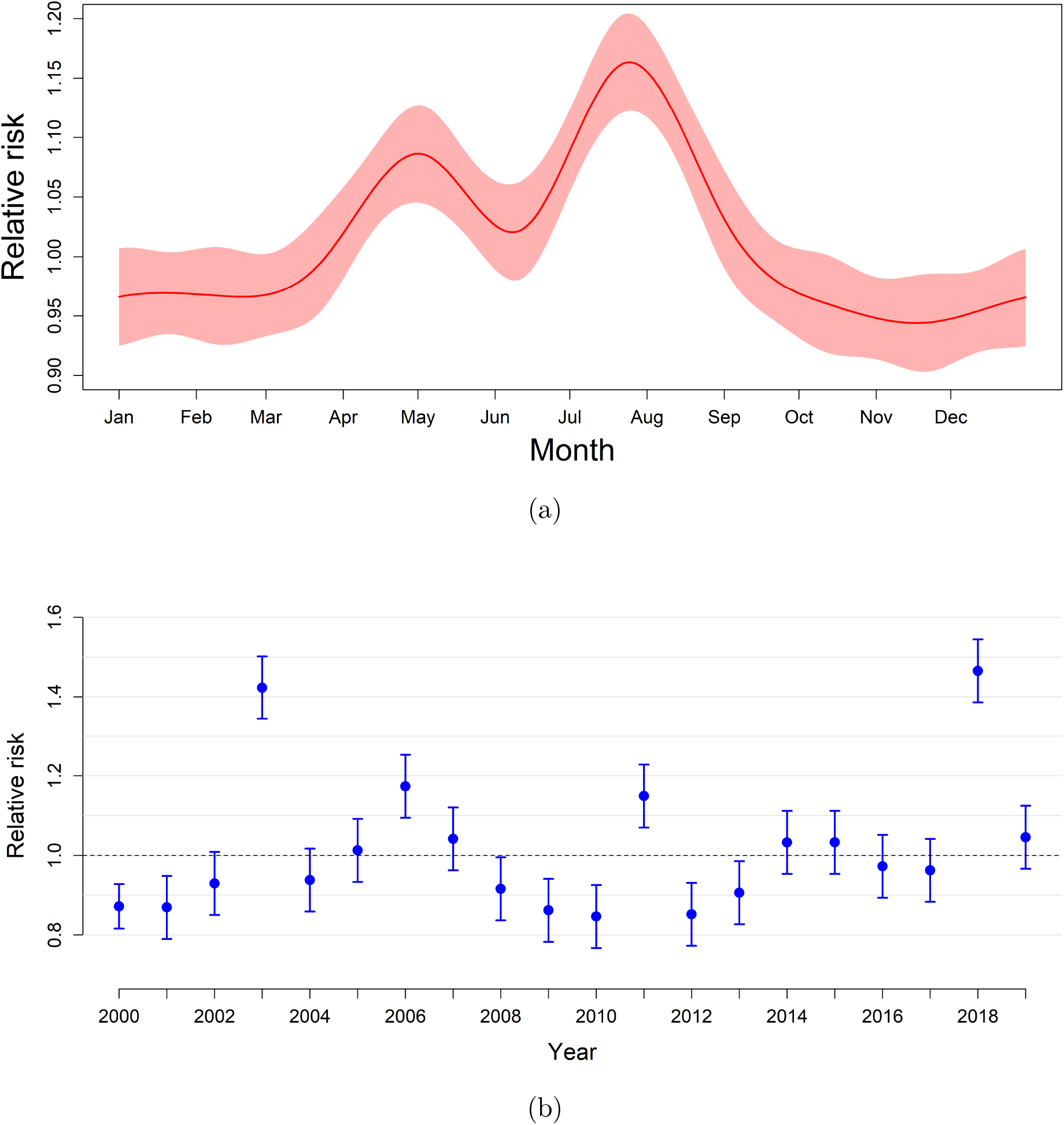
Plots of the meteorological effects that occurred within-year and between-years in 2000–2019.

Seasonal analysis indicated that, if meteorological conditions were made more favorable, expected Legionnaires’ disease incidence would change little from October–March, whereas incidence would increase substantially in late spring and summer (3(a)). Specifically, deviations from the long-term seasonal average revealed a peak effect in May. This suggested that the Legionnaires’ disease incidence in May is sensitive to the preceding weather 60 days prior. Therefore, our results supported the hypothesis that favourable meteorological conditions in March–April created environments conducive to bacterial growth or human exposure, which lead to a raised incidence. A second peak in August reflected sensitivity to favorable conditions in June–July, increasing risk by over 15%. These seasonal peaks possibly result from a combination of elevated temperatures, moderate dewpoint depression, and increased use of water-related facilities, such as air-conditioning systems and spas, which facilitate *Legionella* proliferation and transmission. Late September–early October incidence peaks remained partially unaffected, possibly indicating that deviations from the weather in August–September likely do not affect the resulting incidence of Legionnaires’ disease by late September–early October. This insight may highlight that the peak incidence at this time of the year may be influenced more so by human behaviour.

Next, we analysed the variation between years (Figure 3(b)). The difference in weather-related risk between years was substantial: years such as 2000, 2001, 2012, and 2013 experienced conditions that suppressed risk, whereas 2003 and 2018 presented favorable conditions for Legionnaires’ disease incidence. Although certain years exhibited meteorological conditions favorable for disease spread (e.g., 2003, 2011), incidence remained low in these periods (Figure 4). This underscored that weather was a driver of risk but not a deterministic factor, as chance events and stochastic processes still play a major role in shaping outbreak dynamics. Extending the lag to 0–60 days captured additional variation arising from bacterial replication and environmental persistence 30–60 days prior to infection, emphasizing that antecedent weather conditions can meaningfully shape observed seasonal incidence patterns.

**Figure 4.**
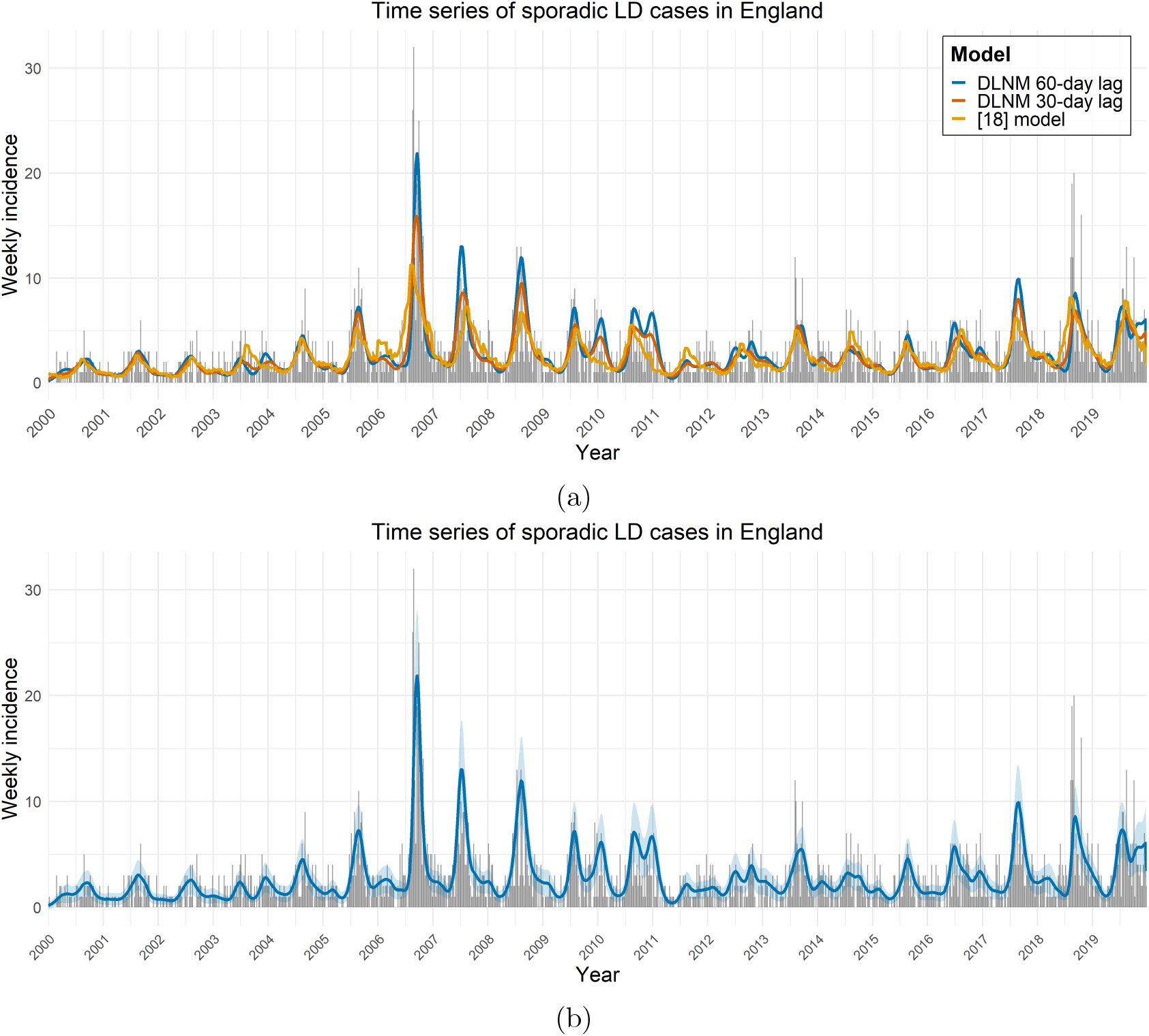
Plots of the nationally aggregated time series of sporadic, community-acquired cases of Legionnaires’ disease. Subfigure (a) shows the GAM fits: the original model [18] in orange, the full model in blue, and the full model with weather limited to 0–30 days prior in red. Subfigure (b) provides the full model with 0–60-days lag, with the respective confidence intervals provided.

### 3.3 Temporal component of the background sporadic model

Time-series analysis provided a framework for understanding the nationally aggregated temporal dynamics of sporadic, community-acquired Legionnaires’ disease incidence. By examining daily case counts over 2000–2019, we captured seasonal patterns, interannual variability, and the lagged effects of environmental factors. This approach disentangled the contributions of weather and other risk factors from stochastic variation, highlighting periods of elevated incidence. We developed a GAM and compared between three formulations: the previous model [18], our model that incorporated 0–60-day weather lags, and our model that only incorporated 0–30-day weather lags. This third model was included to assess the importance of including weather 30–60 days before a case (Figure 4).

The original model [18] successfully captured the overall national temporal trend of weekly Legionnaires’ disease cases (Figure 4(a)); however, it consistently underestimated incidence peaks. This limitation likely arose from the model’s short, one-day lag structure and the absence of region-specific weather covariates, which constrained its ability to account for local environmental conditions affecting bacterial growth and human exposure. Consequently, while the model accurately reflected general national trends, it failed to capture critical variations during periods of elevated risk, highlighting the need for more temporally and spatially nuanced predictors.

To address this limitation, we incorporated 0–30-day, region-specific weather lags, which substantially enhanced the model’s capacity to detect incidence peaks, particularly in 2006 and 2008. By explicitly accounting for meteorological conditions in the month preceding case onset, the model captured environmental drivers influencing both *Legionella* replication and human exposure. This refinement not only improved the characterization of the timing of incidence peaks but also their magnitude, demonstrating that short-term meteorological variability exerted a significant influence on outbreak dynamics.

Building on the 0–30-day lag framework, extending the lag window to 0–60 days further highlighted observed incidence peaks and enabled a more nuanced interpretation of pre-infection environmental conditions. Although central estimates slightly overestimated peaks in 2010 and 2011, the prediction intervals encompassed the observed cases, reflecting residual uncertainty in years with favorable meteorological conditions but low incidence (Figure 4(b)). Collectively, these results indicated that environmental conditions during the period of bacterial growth prior to infection directly influenced subsequent incidence, even when short-term conditions within the infection window remained optimal. This finding underscores the importance of incorporating extended environmental exposure in predictive models to capture the full dynamics of *Legionella* transmission.

While cross-validation (e.g., splitting the time series into training and validation sets) could be used to assess the predictive performance of the GAM, in this work we focus on describing the observed temporal trends rather than making out-of-sample predictions. Consequently, we fit the model to the full dataset to maximize information for estimating seasonal and temporal effects. Future work could extend this framework to include formal predictive validation.

### 3.4 Outbreak-detection modelling

Detecting community-acquired Legionnaires’ disease outbreaks is critical for public health, as it allows identification of locations and sources with high concentrations of the bacteria and taking remedial action. Rapid and accurate detection can inform targeted interventions, reduce exposure, and ultimately prevent cases, saving lives. Therefore, early detection is particularly important given the sporadic and potentially severe nature of Legionnaires’ disease.

We illustrate that outbreak detection varied between our model and the model defined in [18] (Figure 5). When fitting the model using the hierarchical clustering algorithm, the clustering thresholds were selected to balance sensitivity, defined as the proportion of true outbreak cases correctly identified, and specificity, defined as the proportion of sporadic cases correctly excluded. This approach provided a systematic framework for outbreak detection while accounting for spatial variability in reported cases.

**Figure 5.**
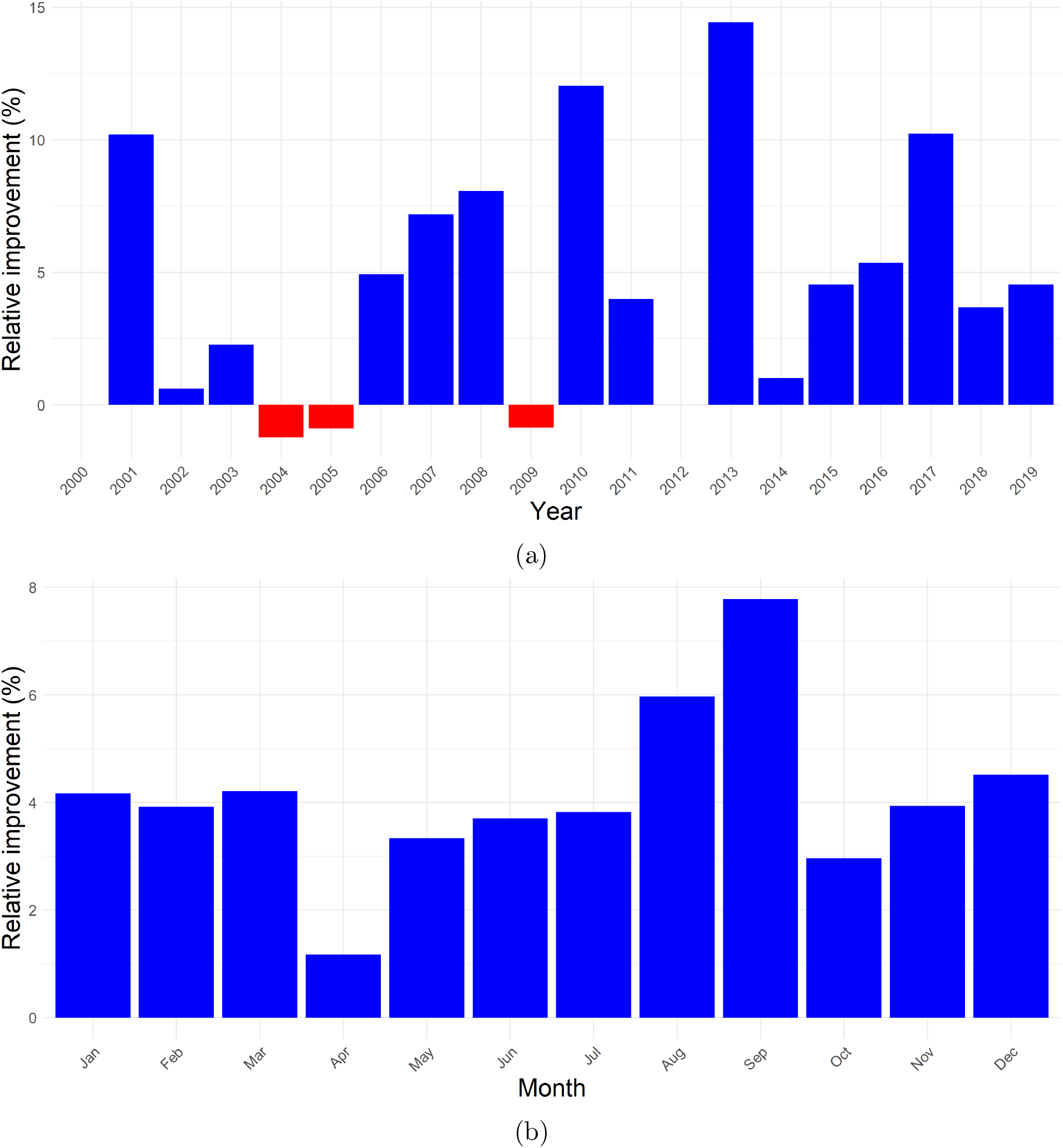
Plots of the relative improvement of our outbreak-detection model compared to [18]. Subfigure represents the relative improvement by year, whereas subfigure (b) represents the relative improvement by month.

Across the 20-year study period, our model outperformed the baseline UKHSA approach in 15 of 20 years, with a mean relative improvement of 6.0% and a maximum improvement of 14.4% in 2013 (Figure 5(a)). In the remaining five years, performance was essentially equivalent in two years or only marginally lower in three years (*≤* 1.0% reduction). In the three years where the baseline model slightly outperformed our approach, no consistent temporal or structural pattern was apparent, suggesting that these differences do not reflect a systematic weakness of the proposed sporadic background model. To assess whether performance gains were concentrated in specific periods of the year, we additionally examined model performance at the monthly level within each year. This analysis indicated that the proposed model improved outbreak detection across all months, with no calendar periods in which it consistently underperformed. Occasional months with slightly reduced performance were sporadic and showed no consistent seasonal or temporal pattern, indicating that improvements were not driven by a narrow subset of high-incidence months.

All evaluations were conducted using grid-based data at a 0.25° latitude–longitude spatial resolution, which inevitably smooths and disperses outbreak signals. At this coarse spatial scale, fine-scale heterogeneity is attenuated, reducing the apparent advantage of models that more effectively capture localized risk. Consequently, the true performance difference between approaches would likely be more pronounced at finer spatial resolutions, such as postcode-level or individual-building data, where the improved ability of our model to distinguish high-risk clusters from sporadic background cases would be more evident. Overall, these results demonstrate that the proposed model consistently provides improved or comparable outbreak detection across multiple seasons, reinforcing its robustness and reliability relative to the existing UKHSA approach [18].

## 4 Discussion

In this study, we developed an integrated spatiotemporal modelling framework for sporadic, community-acquired Legionnaires’ disease that captures fine-scale spatial heterogeneity, extended temporal lags, and deviations in meteorological conditions from local seasonal norms. Unlike many prior analyses, which relied on linear generalized linear model (GLM) assumptions, single-day weather lags, or absolute meteorological values, our approach addresses critical methodological limitations that have hindered consistent interpretation of weather effects. These simplifying assumptions implicitly treat meteorological influences as instantaneous, monotonic, or independent of seasonal context—assumptions that are biologically implausible for *Legionella* ecology. As a consequence, previous studies have reported conflicting directions of association for the same meteorological variables, reflecting the conflation of short-term exposure conditions with longer-term environmental processes. By modelling deviations from expected seasonal baselines and applying a flexible distributed lag nonlinear model (DLNM) framework, we capture the full temporal dynamics of meteorological influence while mitigating collinearity, seasonal confounding, and arbitrary lag selection.

Our framework is structured in three complementary and interdependent stages, each designed to control for distinct sources of variation while informing the others. First, baseline spatial heterogeneity in sporadic Legionnaires’ disease incidence was characterised using a Besag–York–Mollié (BYM2) model incorporating demographic, socioeconomic, and environmental covariates. This approach separates spatially structured from unstructured variation, enabling estimation of persistent regional risk patterns. Elevated risks were concentrated in major urban centres, plausibly reflecting a combination of complex water infrastructure, such as cooling towers and large plumbing systems, and increased opportunities for human exposure. While these mechanisms cannot be directly tested with available data, they are consistent with established epidemiological understanding and provide a rational basis for geographically targeted surveillance. Second, meteorological influences on sporadic incidence were modelled using a DLNM within a matched case–crossover design, allowing flexible estimation of nonlinear and delayed effects over an extended lag window of up to 60 days. Third, spatial risk estimates and DLNM-derived weather effects were integrated into a national time-series negative binomial GAM, with smooth functions capturing long-term trends and random effects accounting for day-of-week variation. This structure allows observed temporal fluctuations to be interpreted as the combined result of persistent spatial susceptibility and transient, weather-driven deviations. Building on these components, we updated the outbreak detection algorithm by replacing the inhomogeneous Poisson assumption with a negative binomial formulation and incorporating refined spatial and temporal risk estimates, thereby improving sensitivity to spatiotemporal clustering while reducing susceptibility to overdispersion.

Analysis of meteorological effects revealed a clear two-stage biological pattern that has not been explicitly resolved in most previous studies. Short-term deviations in temperature and dewpoint depression within the 0–15-day window preceding symptom onset were associated with increased risk, consistent with enhanced bacterial survivability in aerosols and conditions conducive to human exposure during the infection window. In contrast, longer-term deviations were identified occurring approximately 25–50 days prior to onset which require separate investigation to be fully understood; one possibility is that these may reflect distinct processes such as environmental amplification of *Legionella* within potential reservoirs, preceding the more short-term impacts on aerosolization and exposure. This separation of environmental growth and exposure timing provides a mechanistic explanation for the contradictory directions reported in earlier studies, which typically examined only a single temporal window or selected lags based on statistical significance rather than biological plausibility. Our results demonstrate that no single weather variable or short-term lag adequately characterises Legionnaires’ disease risk; instead, incidence emerges from the interaction of cumulative environmental conditions and subsequent exposure opportunities.

The inherent rarity of Legionnaires’ disease further complicates prediction. Infection requires a sequence of stochastic events, including chance encounters with contaminated sources and individual behaviours that are unobserved at the population level. Importantly, this irreducible stochasticity is not a modelling failure but an intrinsic property of sporadic, environmentally mediated infections. Even biologically informed, weather-driven models should therefore be interpreted probabilistically rather than deterministically. While meteorological conditions and population-level susceptibility shape risk landscapes, they are insufficient to predict individual cases with precision, underscoring the need for frameworks that quantify uncertainty rather than attempting exact prediction.

From a public health perspective, the proposed framework improves outbreak detection relative to existing UKHSA methods by more accurately capturing the magnitude and timing of national incidence peaks. Incorporating spatial heterogeneity, extended meteorological lags, and deviations from seasonal norms allows earlier identification of emerging clusters and reduces reliance on fixed baseline assumptions that may obscure environmentally driven signals. By integrating refined spatial risk estimates and weather-sensitive temporal offsets into a hierarchical clustering algorithm, the framework enhances both sensitivity and specificity for detecting potential outbreaks, supporting more timely investigation and intervention. Beyond Legionnaires’ disease, this modelling strategy may be applied to other aerosol transmitted, non-human-to-human pathogens with sporadic incidence patterns, such as *Coxiella burnetii* and *Francisella tularensis*, and non-respiratory *Legionella* infections, and could be adapted to explore disease dynamics under future climate scenarios.

Several limitations should be acknowledged. Higher-order interactions among meteorological variables were not modelled, leaving potential synergistic effects unexplored. Spatial resolution was constrained by available data, and finer-scale exposure information would likely improve both risk estimation and outbreak detection performance. Nonetheless, these limitations do not undermine the central contributions of the framework, which provides a robust foundation for mechanistic interpretation, enhanced surveillance, and hypothesis generation without overstating causal certainty or predictive precision.

In summary, this study demonstrates that integrating spatial structure, extended environmental lag effects, and deviations from seasonal norms yields a more nuanced and biologically coherent understanding of sporadic Legionnaires’ disease dynamics. By explicitly structuring the modelling workflow into three sequential, interdependent stages, we controlled for confounding between spatial, meteorological, and temporal components, producing mutually reinforcing estimates that clarify weather influences and improve outbreak detection. Incidence likely arises from the interplay of environmental conditions, urban infrastructure, human behaviour, and stochastic rare-event processes. By addressing the structural short-comings of prior approaches, this framework provides a flexible and generalizable tool for understanding and monitoring environmentally mediated infectious diseases.

## Data Availability

The individual-level surveillance data used in this study are not publicly available because they contain sensitive health information and were accessed and analysed within secure UK Health Security Agency systems. Access is subject to UK Health Security Agency data-governance requirements and cannot be provided by the authors. The analysis code is publicly available at: https://github.com/NyallJamieson/LD_outbreak_detection

